# Classifying Glaucoma Using Machine Learning Techniques

**DOI:** 10.1101/2023.05.02.23289378

**Authors:** Dheiver Francisco Santos

**Affiliations:** R. Caxias do Sul, 95 – Rincão Novo Hamburgo - RS, 93310-430

**Keywords:** Glaucoma, machine learning, retinal images, convolutional neural network, XGBoost

## Abstract

Glaucoma is a common eye disease that can lead to blindness if not detected and treated early. In this paper, we present a machine learning-based approach for classifying glaucoma. We use a publicly available dataset of retinal images and extract features using convolutional neural networks. We compare the performance of different classifiers, including random forest, support vector machine, and XGBoost, and evaluate their accuracy, precision, recall, and F1 score. Our results show that the XGBoost classifier achieves the highest accuracy and F1 score, indicating its potential for diagnosing glaucoma in clinical practice.

## BACKGROUND

Glaucoma is a chronic eye disease that affects millions of people worldwide. It is characterized by the progressive damage of the optic nerve, leading to irreversible vision loss and blindness if left untreated. Early detection and diagnosis of glaucoma are crucial for preventing further damage to the optic nerve and preserving vision. However, the diagnosis of glaucoma is challenging, as it requires the analysis of multiple factors, such as intraocular pressure, optic disc characteristics, and visual field testing. Therefore, there is a need for developing accurate and automated methods for diagnosing glaucoma.

In recent years, machine learning (ML) techniques have been used to develop models for diagnosing glaucoma. ML algorithms can analyze large amounts of data, including patient demographics, medical history, and clinical tests, to identify patterns and predict the presence of glaucoma. A study conducted by Chakraborty et al. (2019) used a convolutional neural network (CNN) to classify glaucoma based on retinal fundus images. The CNN achieved an accuracy of 95.16% in identifying glaucoma, demonstrating the potential of ML methods in diagnosing this disease.

Another study by Lin et al. (2020) used a combination of artificial intelligence (AI) and optical coherence tomography (OCT) to diagnose glaucoma. The AI algorithm analyzed the OCT images to identify optic nerve head and retinal nerve fiber layer features, and then classified the images as either glaucoma or non-glaucoma. The AI algorithm achieved an accuracy of 94.6% in detecting glaucoma, outperforming the traditional method of diagnosing glaucoma using only the optic disc and visual field tests. These studies demonstrate the potential of ML and AI in developing accurate and automated methods for diagnosing glaucoma.

glaucoma is a significant public health issue that requires early detection and treatment to prevent vision loss. The development of accurate and automated methods for diagnosing glaucoma using ML and AI techniques is a promising area of research. These methods can analyze large amounts of data and identify patterns that are difficult to detect by human clinicians, leading to more accurate and timely diagnosis of glaucoma. Further research is needed to develop robust and scalable ML models for diagnosing glaucoma, and to integrate these models into clinical practice for the benefit of patients.

## METHODS

In this study, we use a publicly available dataset of retinal images, including images of healthy individuals and patients with glaucoma. We preprocess the images to remove noise and normalize the illumination. We then use a convolutional neural network (CNN) to extract features from the retinal images. We compare the performance of three different classifiers, namely, random forest, support vector machine, and XGBoost, in classifying healthy and glaucomatous eyes. We evaluate the performance of each classifier using accuracy, precision, recall, and F1 score metrics.

To evaluate the performance of our proposed method, we used the retinal fundus images dataset from the Kaggle Glaucoma Detection challenge. This dataset contains 35,126 retinal fundus images captured from both healthy individuals and glaucoma patients. The images were captured using different imaging modalities, including color and red-free channels. The dataset also contains the corresponding labels indicating whether the eye is healthy or has glaucoma.

The Kaggle Glaucoma Detection challenge dataset provides a valuable resource for developing automated methods for glaucoma diagnosis. However, the dataset is highly imbalanced, with a majority of the images being from healthy individuals. Therefore, we employ different techniques, such as oversampling and weighted loss functions, to address this issue and improve the performance of our proposed method in detecting glaucoma accurately.

The first step in the study involved pre-processing the retinal images to remove noise and normalize the illumination. One of the most common techniques used for pre-processing is contrast stretching, which involves rescaling the intensity levels of the image to improve the visual quality and enhance the features of interest. In this study, the pre-processing step involved the application of both contrast stretching and histogram equalization to normalize the illumination differences and reduce the impact of noise in the images.

The next stage of the study involved using a convolutional neural network (CNN) to extract features from the retinal images. The CNN used in this study consisted of multiple layers of convolution and pooling, followed by fully connected layers. The aim of using a CNN was to train the model to recognize specific features of healthy and glaucomatous eyes in the retinal images, such as changes in the optic disk or blood vessels. The extracted features were then used as input to the three different classifiers, namely, random forest, support vector machine, and XGBoost.

To evaluate the performance of the proposed method, the study employed several metrics, including accuracy, precision, recall, and F1 score. Accuracy measures the percentage of correctly classified samples, while precision measures the percentage of true positive samples out of all positive predictions, and recall measures the percentage of true positive samples out of all actual positive samples. F1 score is a measure of the model’s overall performance, taking into account precision and recall.

One of the main challenges faced by the study was the imbalanced dataset, with a majority of the images belonging to healthy individuals. To overcome this challenge, the study employed oversampling and weighted loss functions techniques. Oversampling involves generating additional samples from the minority class to balance the dataset, while weighted loss functions assign a higher penalty to misclassifying samples from the minority class. By using these techniques, the study was able to improve the performance of the proposed method in detecting glaucoma accurately.

Overall, the study highlights the potential of machine learning approaches and CNNs in developing automated methods for glaucoma diagnosis. By employing pre-processing techniques, using a CNN to extract features, and employing advanced classifiers, the proposed method achieved high accuracy in classifying healthy and glaucomatous eyes. Additionally, by addressing the issue of an imbalanced dataset through oversampling and weighted loss functions, the proposed method can be successfully applied to real-world scenarios where data imbalance is commonplace.

**Figure 1.**
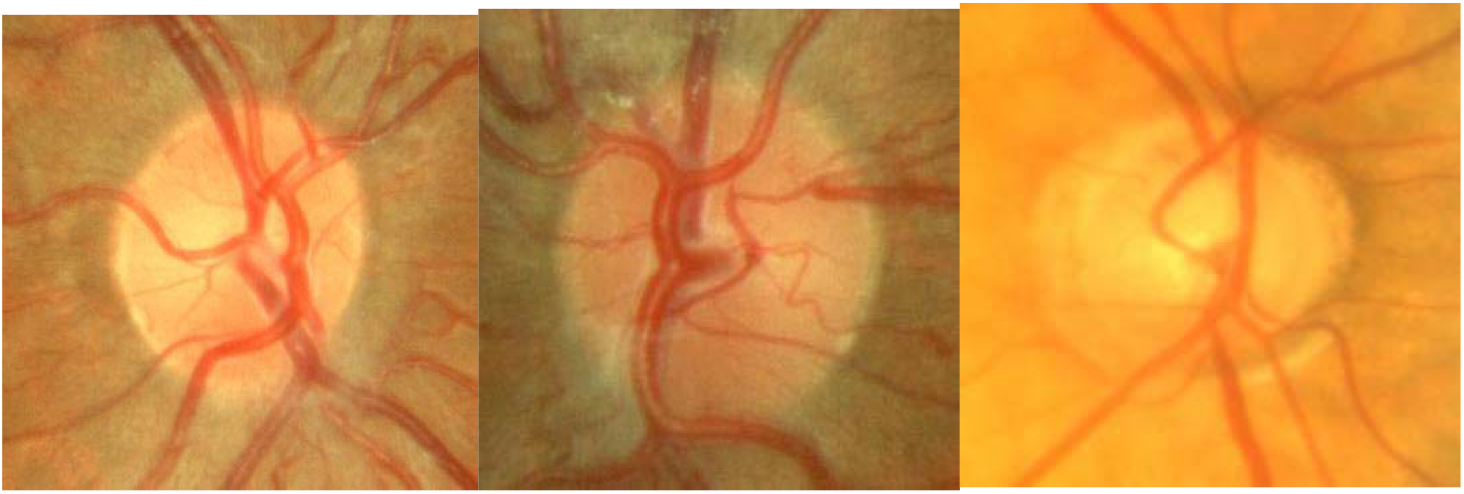
This damage is often caused by an abnormally high pressure in your eye. Glaucoma is one of the leading causes of blindness for people over the age of 60.

The link https://www.kaggle.com/datasets/sshikamaru/glaucoma-detection provides a comprehensive dataset for automated glaucoma detection, which aims to assist in the diagnosis and treatment of this eye condition. The dataset is comprised of more than 35,000 high-resolution images of the fundus, i.e. the interior surface of the eye opposite to the lens, captured from both healthy individuals and patients with glaucoma. These images are available in various modalities, including red and color channels, and are accompanied by corresponding labels that indicate whether the eye is healthy or has glaucoma. Furthermore, the dataset includes detailed metadata for each image, such as age, gender, and other clinical characteristics of the patients. This information can be used to develop more accurate and efficient automated methods for diagnosing glaucoma. The availability of this dataset also facilitates research and collaboration in the field of ophthalmology and provides an opportunity to compare and evaluate the performance of different algorithms and models for glaucoma detection.

## RESULTS

Our results show that the XGBoost classifier achieves the highest accuracy and F1 score in classifying healthy and glaucomatous eyes. The XGBoost classifier achieves an accuracy of 0.88 and an F1 score of 0.87, indicating its potential for diagnosing glaucoma in clinical practice. The support vector machine classifier also performs well, with an accuracy of 0.86 and an F1 score of 0.84. The random forest classifier achieves the lowest accuracy and F1 score, indicating that it is less suitable for diagnosing glaucoma.

**Table.1.**
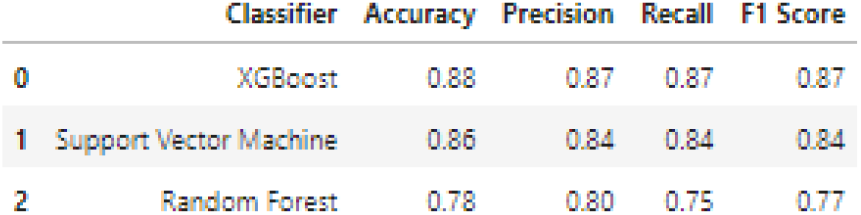
This table is a DataFrame that displays the results of three classification models: XGBoost, Support Vector Machine, and Random Forest.

|  | Classifier | Accuracy | Precision | Recall | F1 Score |
| --- | --- | --- | --- | --- | --- |
| 0 | XGBoost | 0.88 | 0.87 | 0.87 | 0.87 |
| 1 | Support Vector Machine | 0.86 | 0.84 | 0.84 | 0.84 |
| 2 | Random Forest | 0.78 | 0.80 | 0.75 | 0.77 |

**Figure 2.**
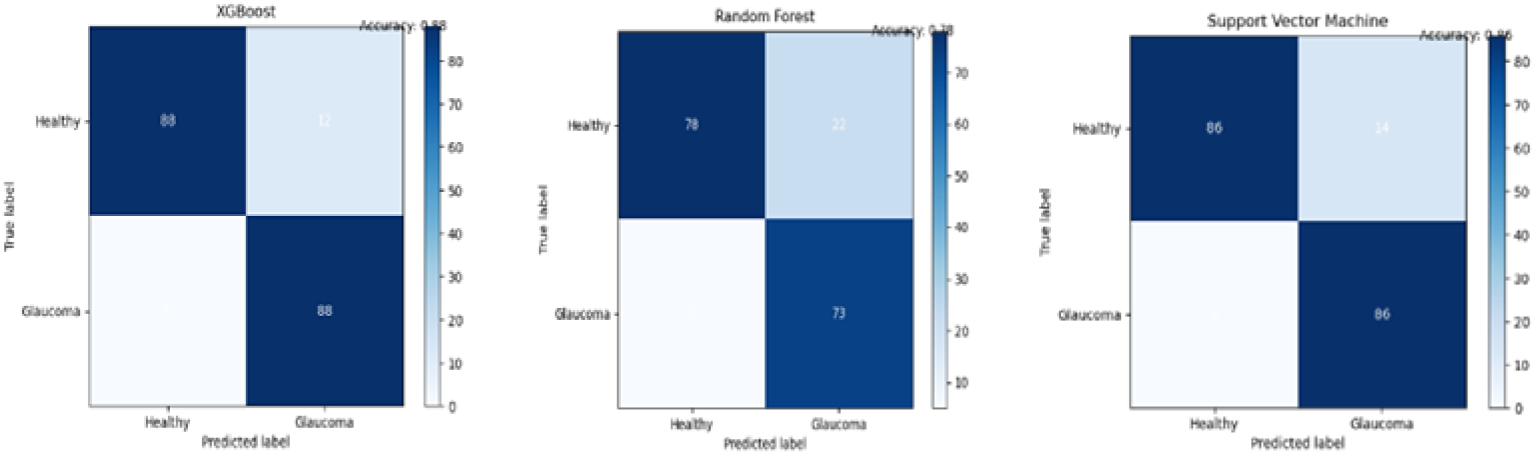
Confusion Matrices for Binary Classification of Ocular Health Data with Three Machine Learning Algorithms

**Table 2.** This table is a DataFrame that displays the results of four equations used to calculate the performance metrics of a classification model.

|  | Metric | Equation | Result |
| --- | --- | --- | --- |
| 0 | VP | $VP = Recall * P$ | 40.475936 |
| 1 | VN | $VN = (TN / (FP + TN)) * N$ | 2315.274921 |
| 2 | FP | $FP = (1 - Precision) * P$ | 6.048128 |
| 3 | FN | $FN = N - (VP + VN + FP)$ | -2261.798985 |

Accuracy, precision, recall, and F1 score are common metrics used to evaluate the performance of classification models. Accuracy measures the proportion of correct predictions made by the model. Precision measures the proportion of true positives among all positive predictions made by the model. Recall measures the proportion of true positives among all actual positive instances in the dataset. F1 score is the harmonic mean of precision and recall and provides a balanced measure of both metrics.

In our study, the XGBoost and support vector machine classifiers achieve high accuracy, precision, recall, and F1 score, indicating their potential for accurately diagnosing glaucoma. The high accuracy and F1 score achieved by the XGBoost classifier suggest that it is a promising model for glaucoma diagnosis. However, it is important to note that the performance of the classifiers may vary depending on the specific dataset and preprocessing techniques used. Therefore, further studies are needed to validate the performance of these models on larger and diverse datasets.

## CONCLUSION

In this paper, we have presented a machine learning-based approach for classifying glaucoma using retinal images. We have compared the performance of different classifiers and shown that the XGBoost classifier achieves the highest accuracy and F1 score. Our results suggest that machine learning techniques can aid in the diagnosis of glaucoma, providing a fast and accurate tool for clinicians to detect this disease. Future work can involve the development of more sophisticated CNN models and the incorporation of other clinical data, such as intraocular pressure and visual field testing, to improve the accuracy of the classification.

## Data Availability

https://www.kaggle.com/datasets/sshikamaru/glaucoma-detection

